# Safe Injection Self-Efficacy is associated with HCV and HIV seropositivity among people who inject drugs in the San Diego-Tijuana border region

**DOI:** 10.1101/2024.05.21.24307696

**Authors:** Katie Bailey, Daniela Abramovitz, Gudelia Rangel, Alicia Harvey-Vera, Carlos F. Vera, Thomas L. Patterson, Jaime Arredondo Sánchez-Lira, Peter J. Davidson, Richard S. Garfein, Laramie R. Smith, Eileen V. Pitpitan, Shira M. Goldenberg, Steffanie A. Strathdee

## Abstract

**Background:** Safe injection self-efficacy (SISE) is negatively associated with injection risk behaviors among people who inject drugs (PWID) but has not been examined in differing risk environments. We compared responses to a validated SISE scale between PWID in San Diego, California and Tijuana, Mexico, and examine correlates of SISE among PWID in Tijuana.

**Methods:** PWID were recruited via street outreach for a longitudinal cohort study from October 2020 – September 2021. We compared SISE scale items by city. Due to low variability in SISE scores among San Diego residents, we restricted analysis of factors associated with SISE to Tijuana residents and identified correlates of SISE scores (low, medium, high) using ordinal logistic regression.

**Results:** Of 474 participants, most were male (74%), Latinx (78%) and Tijuana residents (73%). Mean age was 44. Mean SISE scores among San Diego residents were high (3.46 of 4 maximum) relative to Tijuana residents (mean: 1.93). Among Tijuana residents, White race and having previously resided in San Diego were associated with higher SISE scores. HCV and HIV seropositivity, homelessness, fentanyl use, polysubstance co-injection, and greater injection frequency were associated with lower SISE scores.

**Conclusions:** We found profound inequalities between Tijuana and San Diego SISE, likely attributable to differential risk environments. Associations with fentanyl and polysubstance co-injection, injection frequency, and both HIV and HCV seropositivity suggest that SISE contribute to blood-borne infection transmission risks in Tijuana. SISE reflects an actionable intervention target to reduce injection risk behaviors, but structural interventions are required to intervene upon the risk environment.

## Introduction

People who inject drugs (PWID) are at elevated risk of exposure to blood-borne infections such as Hepatitis C (HCV) and human immunodeficiency virus (HIV), in part due to injection practices such as receptive syringe sharing [1]. An important factor hypothesized to influence disease transmission risk for PWID is self-efficacy to engage in safer injection practices. Self-efficacy is a construct with origins in Bandura’s Social Cognitive Theory (SCT) and refers to a person’s belief in their capacity to complete a given behavior [2].

SCT posits that self-efficacy is one of several internal, cognitive processes that influence behaviors. Cognitive processes are mental functions involving how individuals perceive and interpret information. Moreover, SCT recognizes that behavior is shaped by the dynamic interplay between cognitive processes, behavioral factors, and environmental influences, a concept referred to as reciprocal interactions [2]. Behavioral processes encompass the observable actions undertaken by individuals. Environmental factors are those external to the individual, including physical and social surroundings. In the context of injection drug use (IDU), behavioral factors include the types of drugs PWID use and frequency of use. Environmental factors comprise the risk environment in which drug use occurs, influencing the potential for negative health outcomes related to drug use, such as overdose and sharing of injection equipment that can increase transmission of bloodborne viruses, such as HIV and hepatitis B and C [3].

The general self-efficacy scale (GSE-6) has been shown to be a valid and reliable instrument to measure self-efficacy [4]. Self-efficacy is associated with behavior in a variety of contexts [5], including safer drug injection practices. [6–10] The safe injection self-efficacy (SISE) scale was developed in the multi-site Drug Users Intervention Trial (DUIT) [11] and has been applied among various groups, including PWID in the United States (U.S.) and Mexico [8,9,12]. This research has shown that SISE is an important indicator of IDU risk behaviors. However, studies have not examined how SISE in differing risk environments.

We assessed participant responses to items in the SISE scale among a cohort of PWID recruited through street outreach in San Diego, California and Tijuana, Baja California, Mexico, comparing residents of the two cities. Given San Diego and Tijuana have very different risk environments, particularly with regard to harm reduction service availability, we expected SISE to be higher among San Diego residents. To explore the SCT reciprocal interactions concept, we also assessed associations between cognitive, behavioral, environmental, and health-related factors and low, medium, and high SISE scores (Figure 1).

**Figure 1.**
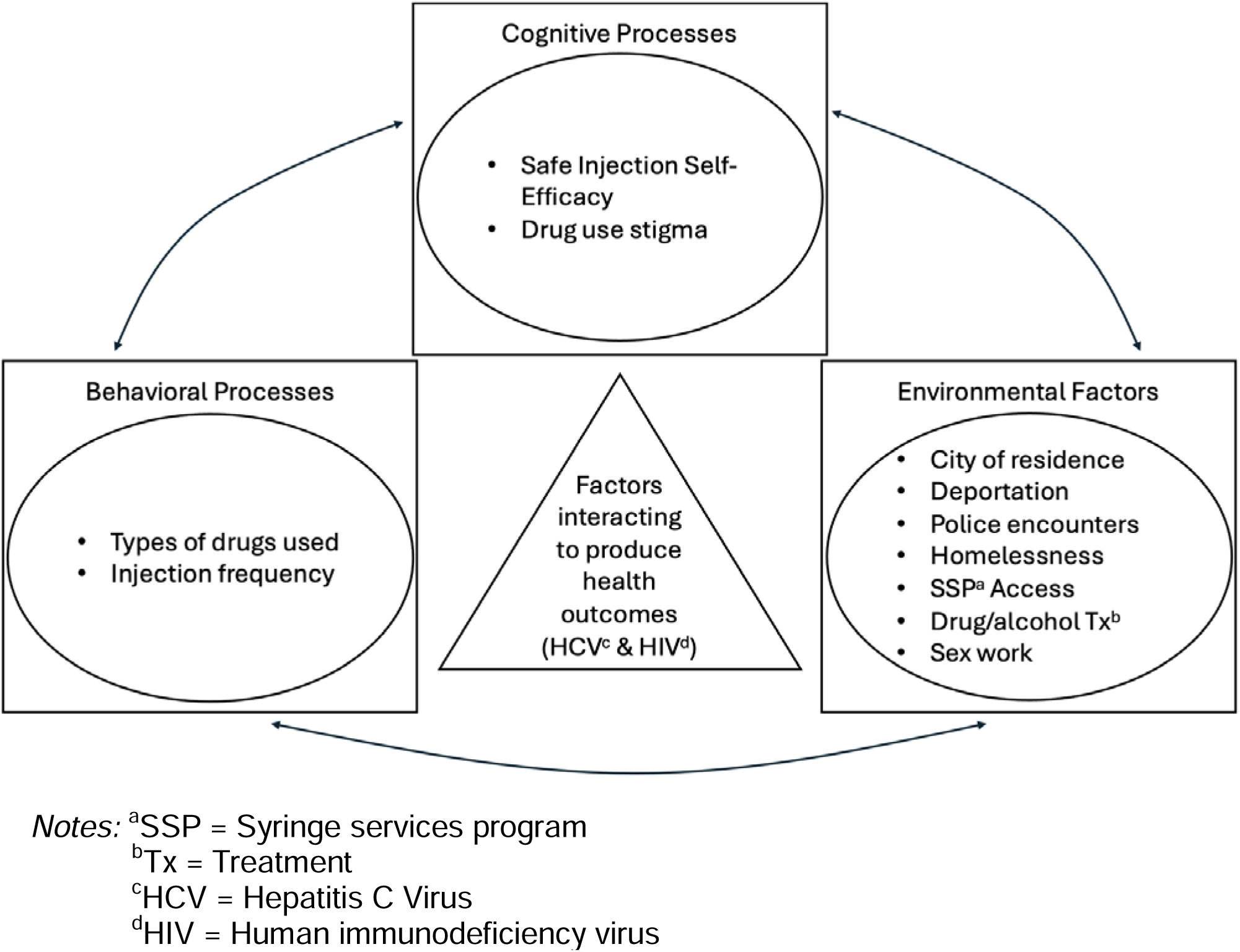
Application of Social Cognitive Theory to assess associations between Safe Injection Self-Efficacy and other cognitive, behavioral, environmental, and health factors.

We explored several hypotheses. Since stigma is associated with injection risk behaviors [13,14], we hypothesized that internalized and anticipated substance use stigma, which are internal cognitive processes related to possessing a socially devalued characteristic [15], would be inversely associated with SISE. Research suggests that fentanyl use [16] (a behavioral process) is associated with more frequent injection among PWID [17]. We therefore hypothesized that fentanyl use would be inversely associated with SISE, as PWID who inject more frequently might feel less self-efficacy to consistently use sterile injection equipment. Additionally, we hypothesized that accessing syringe service programs (SSPs) (an environmental factor) would be associated with greater SISE due to the social and instrumental support (i.e., provision of sterile injection equipment) that SSPs provide. Assessing associations with SISE may help identify PWID who could benefit from risk reduction interventions.

### Setting

The San Diego-Tijuana border is the busiest land border crossing in the U.S. and is considered a high intensity drug trafficking area by the United States (U.S.) Drug Enforcement Administration. [18] The populations of San Diego County and Tijuana are approximately 3.3 million [19] and 2.2 million [20] respectively. Although exact numbers are unknown, there are at least 35,000 PWID in San Diego County [21]. There were 485 people newly diagnosed with HIV in San Diego in 2021, with about 10% of cases attributed to IDU. [22] HCV prevalence in San Diego is 2.1% and an estimated one-third of infections are among PWID [23]. There are an estimated 8,000 – 10,000 PWID in Tijuana [24], with an HCV prevalence of up to 90% [25] and an HIV prevalence of up to 16% among this population [26].

There is overlap in the PWID communities of San Diego and Tijuana, with research suggesting that 15% to 66% of PWID in San Diego cross the border into Mexico to engage in drug use. [27,28] PWID motivations to cross the border from San Diego to Tijuana include perceived accessibility to higher quality and less expensive drugs [28].

## Methods

### Participants and Recruitment

Data were drawn from a prospective cohort study among PWID in the San Diego-Tijuana border region (La Frontera) that was designed to assess drug markets, cross-border mobility and their relationship to incidence of HIV, HCV and overdose, as described elsewhere. [29] Participants were recruited between October 2020 – September 2021 via street outreach. Trained, bilingual field research staff focused recruitment on areas of concentrated drug use in both cities. Inclusion criteria were being 18 years or older, speaking English or Spanish, living in either San Diego or Tijuana, and past-month IDU demonstrated through injection marks. Since an overall goal of La Frontera was to examine cross-border mobility, participants who were San Diego residents were purposively oversampled so that 50% reported engaging in cross-border drug use within the preceding two years.

### Data Collection

Field staff solicited informed consent and conducted computer-assisted interviewer-administered surveys with participants at baseline and every six months thereafter. Surveys were developed in English, translated to Spanish, back-translated to English, and reviewed by bilingual research staff to ensure accuracy. The present study used cross-sectional data from the second follow-up survey which took place between December 2021 – December 2022 because it was the most recently completed follow-up survey in which all participants answered questions pertaining to SISE. Participants were compensated $20 USD for each completed survey. Study protocols were approved by institutional review boards at the University of California San Diego (UCSD) and Xochicalco University.

### Measures

#### Safe Injection Self-Efficacy

The SISE scale measure included six questions asking participants about confidence in their ability to avoid injecting with a needle someone else used or sharing other injection equipment in a variety of scenarios. For example, “I can avoid injecting with a needle someone else used, even if I am injecting with people I know well.” Response options were a Likert scale from 1–4 (1 = absolutely sure I cannot, 2 = pretty sure I can, 3 = pretty sure I cannot, 4 = absolutely sure I can) and scores were averaged for a mean composite score between 1 and 4, where 1 reflects low self-efficacy. The SISE scale has been demonstrated good internal consistency among PWID in the U.S. and Mexico (Cronbach’s alphas = .84 – .94). [8,9,12]

#### Demographics

Participants provided demographic information related to race/ethnicity, sex assigned at birth, gender identity, and age. Race categories were not mutually exclusive. For ease of analysis and interpretation, we created a binary variable indicating whether a participant was White or non-White. Those who indicated a race other than White or reported Latinx, Hispanic, or Mexican ethnicity were classified as non-White.

#### Health measures

Measures of HCV and HIV serostatus were based on serological tests approved for use in the U.S. or Mexico. San Diego participants were tested using Medmira® Miriad combined HIV/HCV immunoassay rapid tests [30]. Positive results were confirmed with a second test using Orasure® HIV and HCV [31]. Tijuana participants were tested with Accurak® HIV and HCV. Positive results were confirmed with Intec® for HIV [32] and Quality® for HCV. Confirmatory testing was conducted at UCSD’s Center for AIDS Research (CFAR) laboratory. Participants were tested for both HCV and HIV infection at baseline and those with a previously negative result were re-tested in follow-up assessments. Pre- and post-test counseling was provided following national guidelines in the U.S. and Mexico. Participants with positive test results were referred for follow-up healthcare.

#### Substance Use Stigma

We included two subscales of the Substance Use Stigma Mechanisms Scale (SU-SMS) that assess experiences of internalized and anticipated drug use stigma [15], and were shown to be valid and reliable among PWUD samples (Cronbach’s alphas = .83 – .94). The internalized stigma and anticipated stigma subscales each include six questions with Likert-scale response options from 1–5 (strongly disagree to strongly agree). Responses to each subscale were used to compute a total mean score. Mean stigma scores can be interpreted as 1 = very low stigma, 2 = low to moderate stigma, 3 = moderate stigma, 4 = moderate to high stigma, 5 = very high stigma. [15]

#### Environmental Factors

A measure of cross-border residence indicated whether participants who indicated their primary city of residence was San Diego at baseline reported their past six-month primary residence as Tijuana in the December 2021 – 2022 follow-up survey. Tijuana residents indicated at baseline if they had ever been deported from the U.S. We measured law enforcement encounters by asking participants if they had been stopped or arrested by police in the past six months. Homelessness was based on participant reports of sleeping always or most often in a shelter or places not meant for habitation, such as the street or in a vehicle. Relatedly, we included a variable indicating how many hours per day participants spent on the street on average. SSP engagement indicated whether participants obtained syringes from an SSP in the prior six months, and whether the SSP was located in San Diego and/or Tijuana. Participants also reported if they had enrolled in a drug or alcohol rehabilitation program in the preceding six months. Finally, we asked whether participants had earned money from sex work in the past six months.

#### Drug Use

We created binary drug use variables indicating whether participants knowingly used each type of substance in the prior six months by any method, regardless of frequency. A polysubstance co-injection variable indicated whether participants reported co-injecting an opioid mixed with a stimulant. Finally, participants estimated an average number of times per day they injected any kind of drug in the prior six months.

### Statistical Analysis

All analyses were conducted using base R [33] and RStudio [34]. First, we compared participant demographic characteristics, HIV and HCV serostatus, drug use stigma scores, environmental factors, drug use, and SISE measures based on their primary city of residence in the prior six months (San Diego vs. Tijuana) using t-tests for continuous variables and Chi-square tests or Fisher’s exact tests for categorical variables.

The outcome of interest was SISE mean scores. Because SISE mean scores were skewed, we created an ordered categorical measure of SISE, which we divided into three ordinal groups based on the range of participant scores (i.e., low, medium, and high). This categorization allowed for more straightforward interpretations of the relationships between independent variables and SISE scores. Additionally, this strategy helped stabilize model predictions by reducing the impact of nonlinearities inherent in the continuous SISE scores.

We used the POLR R package [35] to conduct ordinal logistic regression analyses. First, we assessed univariable associations between the independent variables and the outcome. We included variables that were significant at the p≤0.1 threshold in univariable models in a multivariable, manual forward selection regression modeling process. We added variables to the multivariable model one-by-one prioritizing the lowest p-values and largest effect sizes in univariable models. Only variables that independently maintained significance at the p≤.05 threshold were included in the final model. We evaluated the final model for multicollinearity by examining correlation matrices and variance inflation factors, tested for interactions between covariates and assessed the proportionality of odds assumption using the Brant test [36]. Additionally, we checked for independence of observations and assessed linear relationships between continuous predictors and the log odds of the outcome.

## Results

### Study Sample

The parent study baseline sample consisted of 612 PWID [37]. Eighty-two percent (n=500) of baseline participants completed the December 2021 – 2022 follow-up survey utilized in the present analysis. Missing participants (n=112) were lost due to death (45%, n=50) or otherwise could not be located for follow-up. Over half of deaths (52%, n=25) were confirmed via obituary or local government records and others were reported by next of kin. We removed an additional 26 participants from analysis who indicated they had not injected drugs in the prior six months, for a total analytic sample of 474. Those who were lost to follow-up were more likely to reside in San Diego at baseline and more likely to be White, non-Hispanic (see Appendix, Table 4). Those lost to follow-up were also more likely at baseline to have been experiencing homelessness; have been SSP clients; have been enrolled in drug/alcohol rehabilitation; and have used fentanyl, methamphetamine, or cocaine in the prior six months.

### Safe Injection Self-Efficacy

Overall, the mean SISE score was 2.35 out of 4 (standard deviation [SD]=1.08) and the median score was 2 (interquartile range [IQR]=2). The Cronbach’s alpha for the SISE scale among this sample was 0.99.

### Sample Descriptive Statistics

Among the analytic sample (N=474), 27% (n=129) indicated San Diego was their primary residence in the preceding six months while 73% (n=345) primarily resided in Tijuana (Table 1). This included 154 (33%) participants whose primary residence switched from San Diego to Tijuana during the study period.

**Table 1.** Participant characteristics stratified by prior 6-month city of primary residence in a cohort of people who inject drugs in San Diego, California and Tijuana, Baja California, Mexico, December 2021 – December 2022.

|  | San Diego residents | Tijuana residents | P-value | Overall |
| --- | --- | --- | --- | --- |
| n | 129 | 345 |  | 474 |
| **Demographics** |  |  |  |  |
| Race / Ethnicity (n(%)) <sup>a</sup> |  |  |  |  |
| Non-White Race/Ethnicity (n(%)) | 79 (61.2) | 312 (90.4) | <0.001 | 391 (82.5) |
| Native American (n(%)) | 5 (3.9) | 0 (0.0) | 0.002 | 5 (1.1) |
| Black (n (%)) | 11 (8.5) | 4 (1.2) | <0.001 | 15 (3.2) |
| Asian (n(%)) | 1 (0.8) | 1 (0.3) | 1 | 2 (0.4) |
| Pacific Islander (n(%)) | 2 (1.6) | 0 (0.0) | 0.128 | 2 (0.4) |
| White (n(%)) | 70 (54.3) | 25 (7.2) | <0.001 | 95 (20.0) |
| Other (n(%)) | 47 (36.4) | 188 (54.5) | 0.001 | 235 (49.6) |
| Latinx/Hispanic/Mexican ethnicity (n(%)) | 62 (48.1) | 307 (89.0) | <0.001 | 369 (77.8) |
| Sex assigned at birth (female) (n(%)) | 38 (29.5) | 87 (25.2) | 0.415 | 125 (26.4) |
| Non-binary/trans/other identity (n(%)) | 1 (0.8) | 7 (2.0) | 0.587 | 8 (1.7) |
| Age (mean (SD)) | 43.57 (11.76) | 44.18 (9.61) | 0.565 | 44.01 (10.25) |
| **Safe Injection Self Efficacy (1–4)** |  |  |  |  |
| Mean score (mean(SD)) | 3.46 (0.65) | 1.93 (0.90) | <0.001 | 2.35 (1.08) |
| Median score (median(IQR)) | 4.0 (1.0) | 2.0 (2.0) | <0.001 | 2.0 (2.0) |
| **Health** |  |  |  |  |
| HCV Seropositive (n(%)) | 73 (68.9) | 143 (41.6) | <0.001 | 216 (48.0) |
| HIV Seropositive (n(%)) | 4 (3.8) | 45 (13.1) | 0.012 | 49 (10.9) |
| **Substance Use Stigma** |  |  |  |  |
| Internalized (mean(SD)) | 2.12 (1.12) | 1.87 (0.89) | 0.019 | 1.93 (0.96) |
| Anticipated (mean SD)) | 2.65 (1.26) | 2.81 (1.04) | 0.205 | 2.77 (1.10) |

| **Environmental Factors** |  |  |  |  |
| --- | --- | --- | --- | --- |
| San Diego → Tijuana residence (n(%)) | – | 154 (44.6) | – | 154 (32.5) |
| Deported (reported at baseline) (n(%)) | – | 51 (14.8) | – | 51 (10.8) |
| Stopped/arrested by police<sup>b</sup> (n(%)) | 44 (34.1) | 63 (18.3) | <0.001 | 107 (22.6) |
| Unhoused<sup>b</sup> (n (%)) | 100 (77.5) | 285 (82.6) | 0.258 | 385 (81.2) |
| Avg. hrs./day spent in street<sup>b</sup> (mean(SD)) | 18.0 (8.3) | 14.7 (5.5) | <0.001 | 15.6 (6.5) |
| SSP engagement<sup>b</sup> (n (%)) | 82 (63.6) | 57 (16.5) | <0.001 | 139 (29.3) |
| Drug or alcohol rehabilitation<sup>b</sup> (n(%)) | 30 (23.3) | 10 (2.9) | <0.001 | 40 (8.4) |
| Sex work<sup>b</sup> (n(%)) | 3 (2.3) | 19 (5.5) | 0.222 | 22 (4.6) |
| **Drug Use<sup>b</sup>** |  |  |  |  |
| Fentanyl (n(%)) | 77 (59.7) | 165 (47.8) | 0.028 | 242 (51.1) |
| China white (n(%)) | 7 (5.4) | 70 (20.3) | <0.001 | 77 (16.2) |
| Heroin (n(%)) | 65 (50.4) | 192 (55.7) | 0.357 | 257 (54.2) |
| Rx Opioids (n(%)) | 3 (2.8) | 2 (0.5) | 0.161 | 5 (1.1) |
| Methamphetamines (n(%)) | 102 (79.1) | 194 (56.2) | <0.001 | 296 (62.4) |
| Cocaine (n(%)) | 25 (10.5) | 6 (2.6) | 0.001 | 31 (6.5) |
| Benzodiazepines/Tranquilizers (n(%)) | 10 (7.8) | 76 (22.0) | 0.001 | 86 (18.1) |
| Polysubstance injection (n(%)) | 34 (26.4) | 225 (65.2) | <0.001 | 259 (54.6) |
| Average daily injections (median (IQR)) | 0.3 (2.5) | 2.5 (1.5) | <0.001 | 2.5 (3.7) |
| Notes: <sup>a</sup>Race variables are not mutually exclusive, except for Non-White Race/Ethnicity |  |  |  |  |
| <sup>b</sup>Indicated variables refer to the prior six months |  |  |  |  |

Participants were mostly non-White (83%) and reported Latinx/Hispanic/Mexican ethnicity (78%). Most were assigned male at birth (74%). Participants were an average age of 44 years (SD=10.25). Nearly half (48%) tested HCV-seropositive and 11% tested HIV-seropositive.

Participants reported low to moderate levels of internalized drug use stigma, with an average score of 1.93 out of 5 (SD=0.96), and slightly more moderate levels of anticipated drug use stigma, with an average score of 2.77 (SD=1.10). Cronbach’s alphas for internalized and anticipated stigma subscales were 0.9 and 0.88 respectively.

At baseline, 11% reported having been deported from the U.S to Mexico. Twenty-two percent reported being stopped/arrested in the prior six months. Most (81%) were unhoused in the prior six months, spending an average of 16 hours (SD=6.5) per day on the street. Nearly 30% reported receiving syringes from an SSP in the prior six months, 8% reported being enrolled drug or alcohol rehabilitation program, and 5% had earned money from sex work.

The drugs most commonly used were methamphetamine (62%), followed by heroin (54%), and fentanyl (51%). Just over half reported co-injection. Participants reported a median of 2.5 (IQR = 3.7) injections per day.

### Comparison of San Diego and Tijuana Samples

#### Comparison of Safe Injection Self-Efficacy

The mean and median SISE scores for San Diego residents were significantly higher than that of Tijuana residents (3.46 vs. 1.93 and 4.0 vs. 2.0, p<0.001) (Table 1). For all six scale items, a greater proportion of San Diego participants indicated they were “absolutely sure” or “pretty sure” they could avoid sharing injection equipment in each proposed scenario (Figure 1). Due to low variability in the outcome among San Diego residents, we restricted the ordinal logistic regression analysis to Tijuana residents only.

The SISE mean score distribution among Tijuana residents was right-skewed (i.e., lower SISE scores, see Figure 2). Therefore, we categorized participants into three ordinal groups based on their relative SISE mean scores (low, medium, and high) where low scores were x = 1 (the lowest possible score, N=142), medium scores were 1< x ≤2 (N=89), and high scores were x > 2 (N=114).

**Figure 2.**
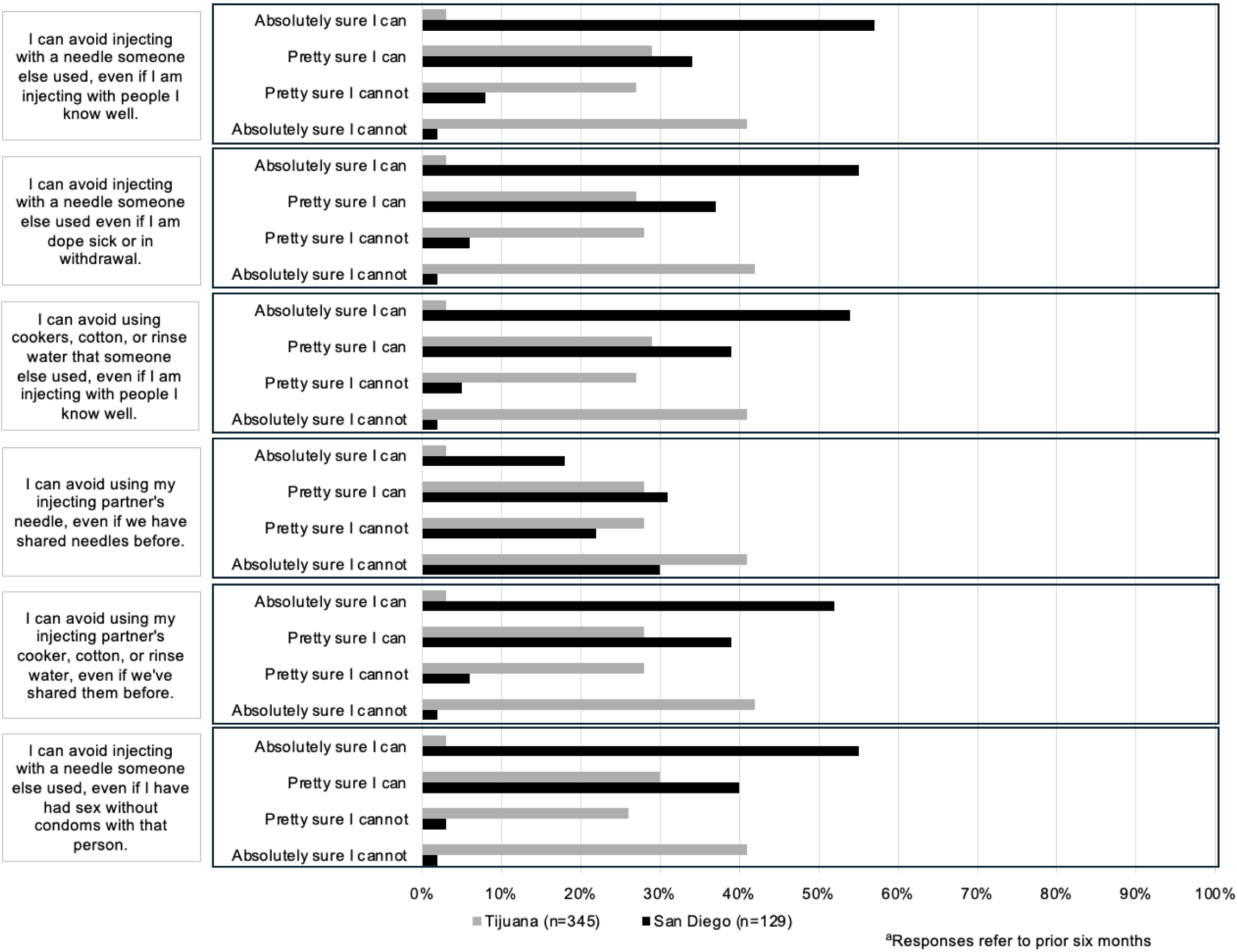
Participant responses to Safe Injection Self-Efficacy questions among a cohort of people who inject drugs in San Diego, California, Mexico, December 2021 – December 2022^a^.

**Figure 3.**
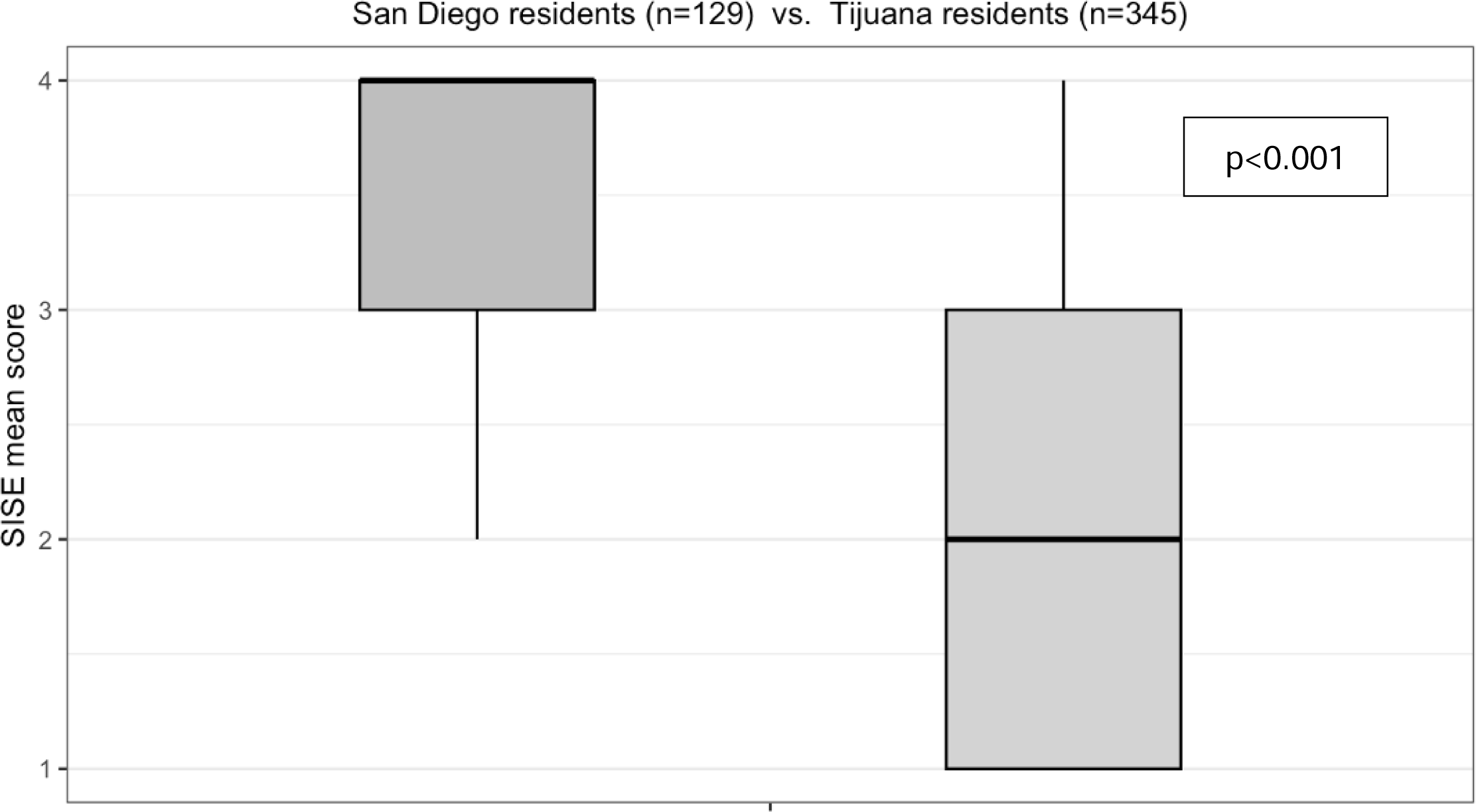
Safe Injection Self-Efficacy (SISE) mean scores by city of residence.

#### Comparison of Descriptive Statistics

Relative to San Diego residents, a higher proportion of Tijuana residents were non-White (90% vs. 61%), with most reporting Latinx/Hispanic/Mexican ethnicity (89%). Compared to Tijuana residents, San Diego residents were more likely to test HCV-seropositive (69% vs. 42%, p<0.001), but less likely to test HIV-seropositive (4% vs. 13%, p=0.012).

While residents of both cities reported low to moderate levels of internalized drug use stigma, mean scores were significantly lower among Tijuana residents compared to San Diego residents (1.87 vs. 2.12, p=0.02), but there was no significant difference in anticipated stigma scores between residents of the two cities.

Compared to Tijuana, a significantly larger proportion of San Diego residents reported past six-month law enforcement encounters (34% versus 18%). Although the difference between cities in terms of homelessness was not statistically significant, San Diego residents indicated they spent a greater number of hours in the street per day relative to Tijuana residents (mean: 18 vs. 15 hours). Compared to Tijuana, SSP engagement was much more common in San Diego (64% vs. 17%) as was drug/alcohol rehab enrollment (23% vs. 3%).

Relative to Tijuana residents, fentanyl and methamphetamine use were more common in San Diego (60% vs. 48% and 80% vs. 56%, respectively). Tijuana residents more commonly reported use of “china white” (a street formulation of fine powder heroin often containing fentanyl) [38] and benzodiazepines or tranquilizers (20% vs. 5% and 22% vs. 8%, respectively). Compared to San Diego residents, Tijuana residents much more commonly reported polysubstance co-injection (65% vs. 26%) and reported a higher number of average daily injections (2.5 vs. 0.3).

### Univariable Associations with Safe Injection Self-Efficacy among Tijuana participants

Due to low variability in SISE scores among San Diego participants, we explored associated factors among the Tijuana resident sample only (n=345). The Cronbach’s alpha for the SISE scale among Tijuana residents was 0.99. In univariable analyses, factors positively associated with SISE scores at the p≤0.1 threshold were female sex, switching primary residence from San Diego to Tijuana during the study period, engaging in sex work, and heroin use (Table 3). Factors negatively associated with SISE were non-White race/ethnicity, HIV and HCV seropositivity, anticipated drug use stigma, ever having been deported, having been stopped/arrested by police, homelessness, more hours spent in the street, SSP engagement, fentanyl use, china white use, benzodiazepine/tranquilizer use, polysubstance co-injection, and reporting a higher number of daily injections during the last six months.

**Table 2.** Categories of Safe Injection Self-Efficacy mean scores among people who inject drugs in Tijuana, Baja California, Mexico, December 2021 – December 2022 (N=345)

| Category level | SISE score range | Mean SISE Score | Size (n) |
| --- | --- | --- | --- |
| High | $x > 2$ | 3.05 | 114 |
| Medium | $1 < x \leq 2$ | 2.00 | 89 |
| Low | $X = 1$ | 1.00 | 142 |

**Table 3.**
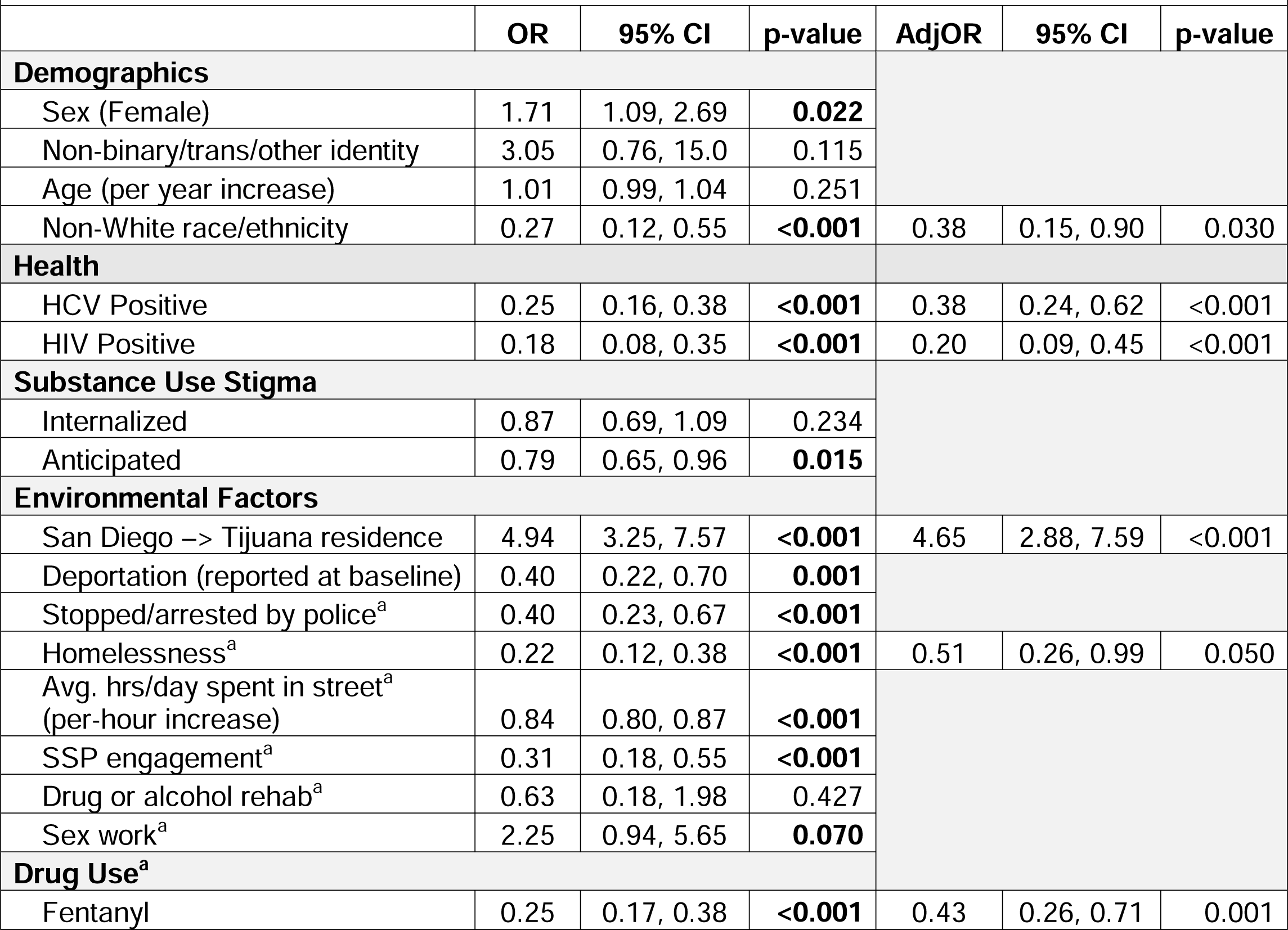
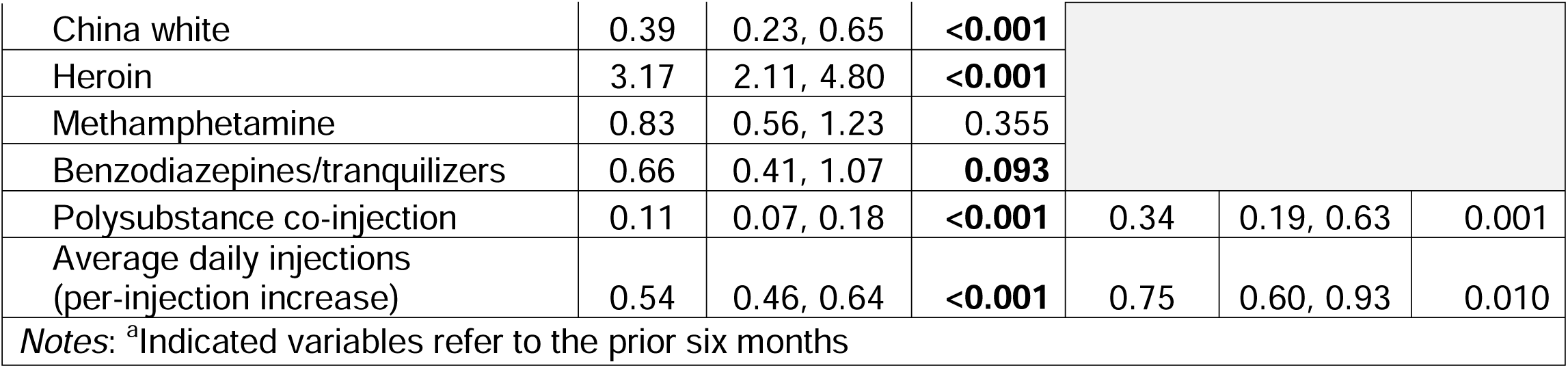
Factors associated with safe injection self-efficacy score (low, medium, high) using univariable and multivariable ordinal logistic regression among people who inject drugs in Tijuana, Baja California, Mexico, December 2021– December 2022 (N=345)

### Multivariable Associations with Safe Injection Self-Efficacy among Tijuana participants

In the final multivariable model, switching residence from San Diego to Tijuana during the study period was the only factor positively associated with SISE (Adjusted Odds Ratio [AdjOR]: 4.65; 95% Confidence Interval [CI]: 2.88–7.59) (Table 3). Non-White race/ethnicity was negatively associated with SISE levels (AdjOR: 0.38; 95% CI: 0.15–0.90), as was HCV-seropositivity (AdjOR: 0.38; 95% CI: 0.24–0.62), HIV-seropositivity (AdjOR: 0.20; 95% CI: 0.09–0.45), and homelessness (AdjOR: 0.51; 95% CI: 0.26–0.99). Three drug use factors were negatively associated with SISE levels: fentanyl use (AdjOR: 0.43; 95% CI: 0.26–0.71), polysubstance co-injection (AdjOR: 0.34; 95% CI: 0.19–0.63), and average number of daily injections (AdjOR: 0.75 for each additional injection per day; 95% CI: 0.60–0.93).

## Discussion

In this binational study of SISE among PWID, we found that participants with primary residence in San Diego at the December 2021 – 2022 follow-up had relatively high SISE sores compared to those residing in Tijuana. Further, Tijuana residents had lower SISE median scores than reported in an earlier study among PWID in Tijuana [8]. Among Tijuana residents, we found a positive association between SISE and prior San Diego residence, and negative associations with non-White race/ethnicity, HIV and HCV seropositivity, homelessness, fentanyl use, polysubstance co-injection, and greater injection frequency. Contrary to our hypothesis, SSP engagement was not significantly associated with higher SISE scores, and the direction of the association was in the opposite direction than anticipated. This relationship may be explained by research suggesting that SSP participants may engage in more high-risk drug use relative to those who are not engaged with SSPs [39].

Although SISE is an individual-level psychological construct, it may interact with and be influenced by environmental factors as described by SCT. There were key differences in access to harm reduction services between San Diego and Tijuana that likely explain some of the relative differences in SISE scores between participants in these cities. Amidst the escalating severity of the addiction and overdose crises in the U.S., there has been growing political and government support for harm reduction initiatives at the national, state, and local levels [40–42], notwithstanding historical resistance that continues to stymie progress [43,44]. In recent years this has translated to an increase in harm reduction service provision in San Diego, including new SSP providers.

Conversely, harm reduction services are more limited in Tijuana. Despite supportive harm reduction policies introduced in Mexico near the turn of the millennium, including partial decriminalization of drug possession [45], scholars have identified a current lack of political will to comprehensively confront addiction and overdose issues prevalent in Mexico-U.S. border cities [46]. Rescinded global [24] and federal government funding to civil society organizations [47] has had a negative impact on SSP engagement and has led to increased receptive syringe sharing among PWID in Tijuana [24]. Additionally, there are significant barriers to syringe access via pharmacies in Tijuana [48]. The threat of violence related to organized crime may also impede access to SSP services among PWID in Tijuana [49]. These key differences in harm reduction service provision and access between San Diego and Tijuana are thought to be key drivers in the differential risk environments of the two cities [3]. Our findings that White race and prior residence in San Diego were positively associated with SISE among Tijuana resident-participants are likely reflections of these distinct differences. Accordingly, those who had access to harm reduction services in San Diego, despite primarily residing in Tijuana, perceived greater self-efficacy to employ safe injection practices.

Our finding that HCV and HIV serostatus was negatively associated with SISE has important ramifications for public health. Prior research has shown that behavioral interventions that enhance SISE can reduce the risk of acquiring blood-borne infections among female sex workers who engage in IDU in Mexican border cities [50]. Motivational interviewing [51] and risk education interventions [52] have also been shown to reduce injection risk behaviors. While lower SISE is associated with future injection risk behaviors, it is also possible that an individual’s sense of SISE is deflated following a positive HCV or HIV test. As such, a positive HCV or HIV test may be an important time to implement interventions to bolster SISE. Efforts to scale up these individual-level interventions to improve SISE among PWID in Tijuana are warranted. However, such efforts may be undermined by ongoing structural barriers in Tijuana that require intervention at a policy level. Indeed, proposed legislation in the U.S. and Mexico to further criminalize fentanyl use would likely intensify health risks for PWID [53].

Our findings suggest the need to increase access to rapid point-of-care HCV and HIV testing among PWID in Tijuana [54,55]. HCV testing, diagnosis, and counseling alone can reduce injection risk behaviors among PWID, ultimately reducing further disease transmission [56]. Importantly, HCV can be treated and cured, although barriers to treatment among PWID persist [57]. More frequent testing among PWID is key for reaching HCV and HIV elimination targets set by the World Health Organization, to which Mexico has committed [58]. Recent research among PWID in the San Diego-Tijuana border region suggests the high potential for HIV self-testing [59], particularly for those who do not regularly access health services. HCV self-testing may also be a feasible way to increase uptake among PWID [60], although these testing platforms are not yet approved in either country. Harm reduction services that prioritize destigmatizing practices are important intervention points for facilitating HCV and HIV testing and treatment connections in this population [57]. HIV prevention medications including pre-exposure prophylaxis (PrEP) and post-exposure prophylaxis (PEP) are also vital prevention measures, although identifying strategies to increase awareness and uptake among PWID is needed [61,62].

Illicitly manufactured fentanyl has a shorter half-life relative to heroin and other opioids [63], and is associated with increased injection frequency among PWID to avert withdrawal symptoms and maintain a prolonged effect [64]. As such, it is logical that this behavioral factor and other indicators of heightened injection frequency, such as polysubstance co-injection, were negatively associated with SISE. Importantly, recent research by members of our team found fentanyl use to be an independent predictor of HCV seroconversion [65]. Our study suggests SISE could be a mediator of this relationship through its potential impact on injection risk behaviors, which warrants further study. Improved access to a sufficient supply of sterile syringes is needed for PWID who inject frequently. Additionally, increasing access to medications for opioid use disorder (MOUD) treatment with nuanced dosing approaches tailored to persons with fentanyl addiction can reduce injection frequency [66]. However, there is currently a shortage of affordable MOUD services in Tijuana [67], intensified by the closing of the only methadone manufacturer in Mexico in 2023 [68]. Finally, the expansion of safe smoking supplies may also reduce injection frequency and subsequently enhance SISE if PWID smoke instead of inject drugs [69].

In our study, homelessness was associated with lower levels of SISE, which is an environmental factor consistently found to be associated with riskier drug use behaviors among people who use drugs (PWUD) [70,71]. Homelessness is related to other environmental factors that may influence cognitive processes like self-efficacy. For example, people experiencing homelessness have limited capacity to carry sterile injection equipment and access harm reduction services in light of regular displacement, police harassment, violence, and incarceration [72–74]. Involuntary displacement of PWUD experiencing homelessness is a common practice that exacerbates health problems [73]. Interventions to expand access to low-barrier, stable housing may help reduce injection risk behaviors [75] and improve many other health outcomes for this population [76].

## Limitations

Participants recruited for the present study were not randomly sampled and represent a particularly marginalized community of PWID in the San Diego-Tijuana border region; therefore, our findings may not generalize to other cities or rural populations. Additionally, participants residing in San Diego at baseline were oversampled to ensure representation of cross-border drug use. As such, findings may not represent the broader population of PWUD in the region.

Some measures used in analysis might underestimate risk factor estimates. The deportation measure only reflected those who indicated at baseline that they had been deported; it is unknown if any participants who switched residence from San Diego to Tijuana during the study period were deported. Additionally, our drug use measures were based on self-report, yet participants may have unintentionally or unknowingly used some substances (e.g., fentanyl). Our SSP measure was based on having received syringes from an SSP in the prior six months but may underestimate SSP engagement for other service needs, and does not account for secondary syringe distribution from SSPs. The SISE categories used in the present study were created based on the distribution of SISE scores in the Tijuana sample. These categories should not be directly compared to SISE score categories in other studies.

Our study findings are subject to survivorship and attrition bias. PWID lost to follow-up had indications of both higher and lower risk based on several structural and drug-use factors. Finally, our study utilized cross-sectional data, precluding causal and temporal inferences between independent variables and SISE.

## Conclusions

Our study suggests that SISE can vary significantly by drug use risk environment. Further, our findings highlight the potential for SISE interventions to impact HCV and HIV transmission risk among PWID, particularly for those who inject more frequently.

## Data Availability

All data produced in the present study are available upon reasonable request to the authors.

## Appendix

**Table 4.** Study retention by baseline characteristics^a^.

| **Table 4. Study retention by baseline characteristics<sup>a</sup>** |  |  |  |  |
| --- | --- | --- | --- | --- |
|  | Not lost to follow-up | Lost to follow-up | p-value | Overall |
| n | 474 | 112 |  | 586 |
| Residence city (Tijuana) (n(%)) | 191 (40.3) | 9 (8.0) | <0.001 | 200 (34.1) |
| Non-White Race/Ethnicity (n(%)) | 391 (82.5) | 75 (67.0) | <0.001 | 466 (79.5) |
| Hispanic Ethnicity (n(%)) | 369 (77.8) | 54 (48.2) | <0.001 | 430 (72.2) |
| Sex assigned at birth (female) (n(%)) | 125 (26.4) | 28 (25.0) | 0.859 | 153 (26.1) |
| Non-binary/trans/other identity (n(%)) | 6 (1.3) | 0 (0.0) | 0.500 | 6 (1.0) |
| Age (mean(SD)) | 43.1 (10.3) | 43.0 (12.6) | 0.939 | 43.1 (10.7) |
| HCV Seropositive (n(%)) | 170 (35.9) | 51 (45.5) | 0.073 | 221 (37.7) |
| HIV Seropositive (n(%)) | 42 (8.9) | 3 (2.7) | 0.044 | 45 (7.7) |
| Ever deported | 51 (10.8) | 3 (2.7) | 0.013 | 54 (9.2) |
| Stopped/arrested by police<sup>b</sup> (n(%)) | 161 (34.0) | 25 (22.3) | 0.023 | 186 (31.7) |
| Unhoused<sup>b</sup> | 280 (59.1) | 85 (75.9) | 0.001 | 365 (62.3) |
| SSP engagement<sup>b</sup> (n(%)) | 151 (31.9) | 58 (51.8) | <0.001 | 209 (35.7) |
| Drug or alcohol rehabilitation<sup>b</sup> (n(%)) | 12 (2.5) | 10 (8.9) | 0.003 | 22 (3.8) |
| Sex work<sup>b</sup> (n(%)) | 48 (10.1) | 3 (2.7) | 0.020 | 51 (8.7) |
| Fentanyl<sup>b</sup> (n(%)) | 146 (30.8) | 46 (41.1) | 0.049 | 192 (32.8) |
| China white<sup>b</sup> (n(%)) | 92 (19.4) | 18 (16.1) | 0.497 | 110 (18.8) |
| Heroin<sup>b</sup> (n(%)) | 375 (79.1) | 88 (78.6) | 1.000 | 463 (79.0) |
| Rx Opioids<sup>b</sup> (n(%)) | 40 (8.4) | 9 (8.0) | 1.000 | 49 (8.4) |
| Methamphetamine<sup>b</sup> (n(%)) | 343 (72.4) | 96 (85.7) | 0.005 | 439 (74.9) |
| Cocaine<sup>b</sup> (n(%)) | 47 (9.9) | 24 (21.4) | 0.001 | 71 (12.1) |
| Benzodiazepines/Tranquilizers<sup>b</sup> (n(%)) | 141 (29.7) | 11 (9.8) | <0.001 | 152 (25.9) |
| Polysubstance co-injection<sup>b</sup> (n(%)) | 282 (59.5) | 54 (48.2) | 0.039 | 336 (57.3) |
| Average daily injections<sup>b</sup> (median (IQR)) | 2.5 (3.3) | 2.5 (3.7) | 0.221 | 2.5 (3.3) |
| *Notes:* <sup>a</sup>This table does not include participants who were removed from analysis because they no longer injected drugs at the December 2021 – December 2022 follow-up<br><sup>b</sup>Indicated variables refer to the prior six months |  |  |  |  |

